# The Polygenic Score Catalog: an open database for reproducibility and systematic evaluation

**DOI:** 10.1101/2020.05.20.20108217

**Authors:** Samuel A. Lambert, Laurent Gil, Simon Jupp, Scott Ritchie, Yu Xu, Annalisa Buniello, Gad Abraham, Michael Chapman, Helen Parkinson, John Danesh, Jacqueline A. L. MacArthur, Michael Inouye

**Author notes:** Corresponding authors: MI, SAL, JALM.

## Abstract

Polygenic [risk] scores (PGS) can enhance prediction and understanding of common diseases and traits. However, the reproducibility of PGS and their subsequent applications in biological and clinical research have been hindered by several factors, including: inadequate and incomplete reporting of PGS development, heterogeneity in evaluation techniques, and inconsistent access to, and distribution of, the information necessary to calculate the scores themselves. To address this we present the PGS Catalog (www.PGSCatalog.org), an open resource for polygenic scores. The PGS Catalog currently contains 192 published PGS from 78 publications for 86 diverse traits, including diabetes, cardiovascular diseases, neurological disorders, cancers, as well as traits like BMI and blood lipids. Each PGS is annotated with metadata required for reproducibility as well as accurate application in independent studies. Using the PGS Catalog, we demonstrate that multiple PGS can be systematically evaluated to generate comparable performance metrics. The PGS Catalog has capabilities for user deposition, expert curation and programmatic access, thus providing the community with an open platform for polygenic score research and translation.

## Main Text

By aggregating the effects of many genetic variants into a single number, polygenic scores (PGS) have emerged as a method to predict an individual’s genetic predisposition for a phenotype^1–4^. Early studies indicated that combining allelic counts of Genome-wide Association Study (GWAS)-significant variants in individuals was predictive of the phenotype^5–8^. Owing to larger and more powerful GWAS, more recent PGS typically comprise hundreds-to-millions of trait-associated genetic variants which are combined using a weighted sum of allele dosages multiplied by their corresponding effect sizes.

Many PGS have been developed and demonstrated to be predictive of common traits (e.g. body mass index [BMI]^9^, blood lipids^10^, educational attainment^11^). Similarly, PGS for various diseases have been shown to be predictive of disease incidence, defining marked increases in risk over the lifecourse or at earlier ages for those individuals with high PGS (e.g. coronary artery disease [CAD]^12,13^, breast cancer^14^, schizophrenia^15^). Existing risk prediction models using traditional risk factors can be improved by incorporating PGS^12,16,17^. In some cases PGS may be the most informative risk factor in pre-symptomatic individuals^1,18^, and for some diseases independent of a family history of the condition^19–22^. Other potential clinical uses of PGS include predicting prognosis, aetiology and disease subtypes; stratification of patients according to therapeutic benefit and identification of new disease biomarkers and drug targets. Given their multiple applications, a large number of PGS have been developed, with over 900 articles indexed in PubMed since 2009^23^.

There is widespread variability in PGS research, even with regard to nomenclature: they can be referred to as genetic or genomic scores, and as polygenic risk scores (PRS) or genomic risk scores (GRS) if they predict a discrete phenotype (such as a disease)^24^. There are also many approaches to derive PGS using individual level genotype data or GWAS summary statistics^25^. The goals of most computational methods are to select the most predictive set of variants in the score, and to adjust their weights to maximise predictive capacity and account for linkage disequilibrium (LD) between variants.

### The need for an open resource for polygenic scores

Multiple barriers inhibit progress in PGS research and the translation of PGS into healthcare settings. Lack of best practices and standards, particularly with regard to PGS reporting, are major issues identified by our group and others^24,26^. Reproducibility has been hampered by underreporting of key PGS information; ~33% of 165 papers we reviewed during our curation efforts did not have adequate variant information (e.g. chromosomal location, effect allele and weight) to calculate the PGS for new samples.

Apart from information necessary for PGS calculation, a complete understanding of a score’s ability to accurately predict its target trait (also known as analytic validity) is necessary to help evaluate clinical utility and enable other applications of PGS. However, the performance reported for existing PGS are conditional on study design, participant demographics, case definitions, and covariates adjusted for in the original study’s models. While there are few direct evaluations of PGS, benchmarking of multiple PGS for the same trait in external data provides the comparable performance metrics needed to decide which PGS offers the best performance for a particular task and how this varies when important factors change, such as ancestry^27^. Since PGS are based on data and cohorts of largely European ancestries, there is a well-characterised underperformance of PGS when applied to non-European individuals, thus the transferability of PGS performance is a particularly important challenge^28–30^.

Here, we present the Polygenic Score Catalog (PGS Catalog; www.PGSCatalog.org): an open resource of published PGS annotated with relevant metadata required for accurate application and evaluation. The PGS Catalog promotes PGS reproducibility by providing a venue to annotate and distribute scores according to current exemplar reporting standards. As such, it enables users to re-use and evaluate polygenic scores, thus firmly establishing their predictive ability and facilitating studies to investigate clinical utility.

### Development of the PGS Catalog

The aim of the PGS Catalog is to index and distribute the key aspects of each PGS (underlying variants, results, and experimental design) in a standardised representation, in order to facilitate evaluations of analytic validity. To maximise usability, the data representation and database were designed to be findable, accessible, interoperable, and reusable (FAIR) according to established principles for scientific data management (Supplemental Table 1)^31^.

To define the key information that would need to be captured in the PGS Catalog we undertook an initial literature review of 27 highly-cited publications that developed PGS for the following traits and diseases based on their potential clinical utility and public health burden of disease: coronary artery disease (CAD), diabetes (types 1 and 2), obesity / body mass index (BMI), breast cancer, prostate cancer and Alzheimer’s disease. During our review we took note of how PGSs were described, how they differed between studies and traits, as well as the most common study designs and PGS evaluation scenarios. To capture common aspects of PGS studies we built upon the NHGRI-EBI GWAS Catalog’s established frameworks to catalog published data from genomic studies, using established conventions for representing sample ancestry ^32^, variant, and trait information ^33^. Using our survey and established frameworks we defined four major data objects: **Scores**, **Samples**, **Performance Metrics**, and **Publications** (Box 1, Supplemental Table 2). These objects describe the common PGS development and evaluation processes (Figure 1A), and can be used to capture the detailed data elements necessary to evaluate PGS development and performance.

**Figure 1.**
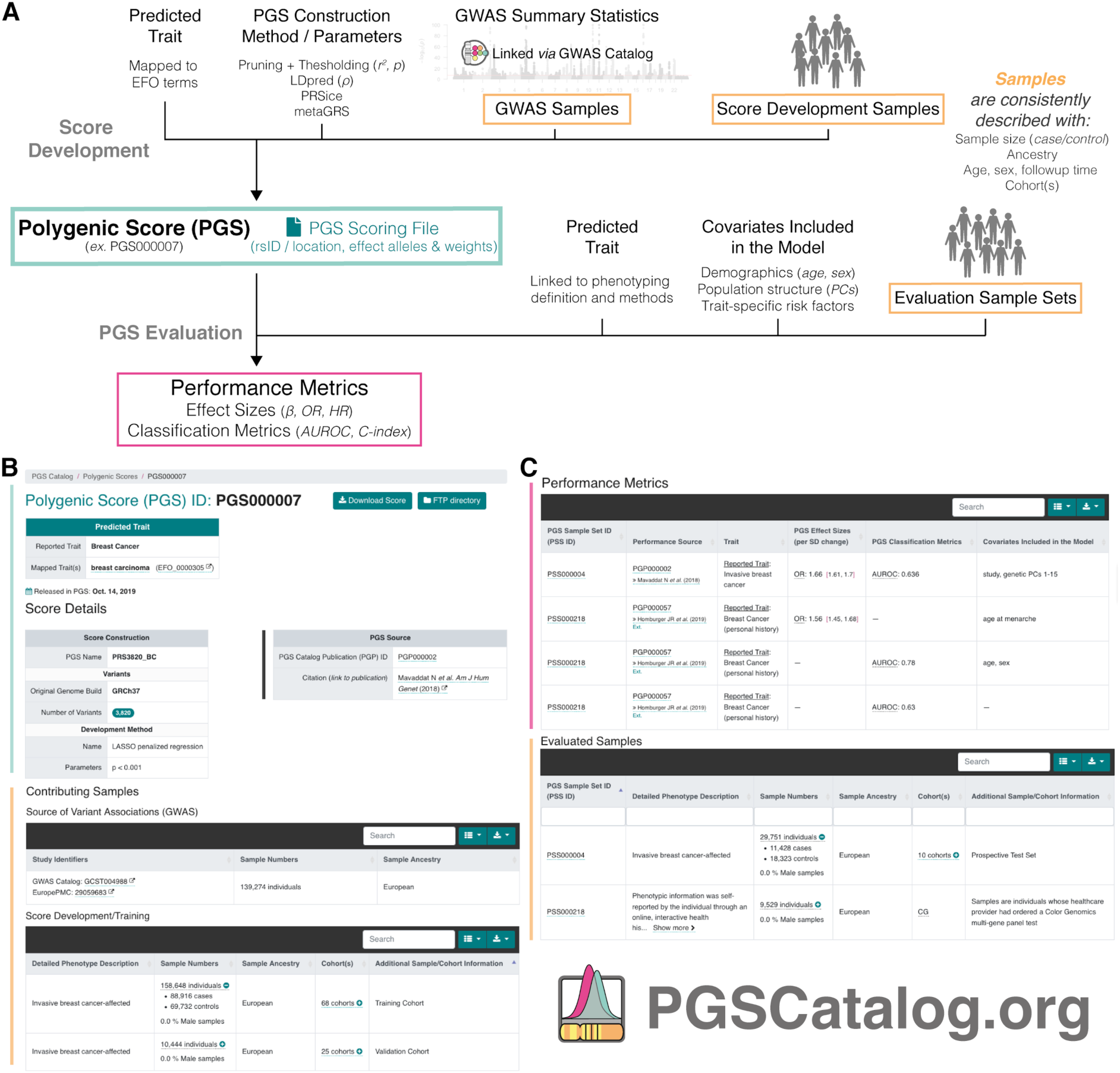
Common aspects of PGS analyses that are captured and displayed in the PGS Catalog. (**A**) PGS analyses can broadly be described in two stages: determining the set of variants and weights that will predict a trait of interest (Score Development), and an evaluation of how predictive the PGS is in an external set of samples (PGS Evaluation). Major data items (Box 1) that can be queried and browsed in the PGS Catalog are highlighted as coloured boxes, and linked to metadata items that are recorded. (**B-C**) Example of how PGS metadata is displayed for each score on PGSCatalog.org (example score PGS00007 ^14^). Sections are highlighted with coloured bars corresponding to the data objects they display in **A**.

To ensure that the PGS Catalog contains the information necessary to describe and evaluate PGS, we collaborated with the ClinGen Complex Disease Working group^34^, composed of experts in epidemiology, statistics, implementation science and the actionability of genetic results, as well as those with disease-domain specific knowledge and interests in PRS application. Together we developed the Polygenic Risk Score Reporting Standards (PRS-RS)^24^, a joint statement describing a set of reporting items that should be described in studies developing and evaluating PRS. The PGS Catalog captures the data required by the PRS-RS to assess PGS validity, while also being flexible enough to capture multiple different study designs and evaluation scenarios in a structured database. The PGS Catalog therefore provides a venue to index PGS analyses and maximize uptake of these reporting standards.

#### Box 1

##### Description of the PGS Catalog objects and metadata

*(Field-by-field reporting items are available in Supplemental Table 2)*

**Scores** (e.g. PGS/PRS/GRS) are the main data object-type in the PGS Catalog, linked to all other objects internally and can be cited or externally linked to by its persistent identifier (e.g. PGS000018). Each PGS is annotated with information about the phenotype it predicts (Reported Trait), and mapped to Experimental Factor Ontology (EFO) terms^35,36^ to consistently annotate related scores and facilitate data linkage and search. Score development details, including computational algorithms and parameters are recorded for each score. The GWAS summary statistics used to derive the model, if any, are linked as **Sample** objects and further linked to the GWAS Catalog if applicable^33^; any other datasets used for training are also linked as **Sample** objects. Each PGS has a **PGS Scoring File**, a flat text file in a consistent format (Supplemental Note 1) which contains the variant-level information necessary to calculate the score on new data (minimally the genome build, rsID or chromosomal positions, effect alleles and their weights).

**Samples** are described with detailed information to enable the interpretation and assessment of the validity of a PGS. Sample size (stratified by cases and controls if dichotomous) and participant ancestry are described using frameworks identical to the GWAS Catalog - this enables the systematic tracking of participant diversity in PGS^37^. To facilitate reproducible analyses, phenotyping descriptions (e.g. case definition, ICD-9/10 codes, measurement methods), the sex distribution, and the distributions of participant ages and follow-up times for prospective study designs can also be recorded. To ensure that PGS are not evaluated on individuals who contributed to the original GWAS or PGS training cohorts, **Samples** can be annotated with existing cohort names^38^. Groups of **Samples** used to evaluate PGS are given a **Sample Set** (PSS) ID.

**Performance Metrics** assess the validity of a PGS in a Sample Set, independent of the samples used for score development. Common metrics include standardised effect sizes (odds/hazard ratios [OR/HR], and regression coefficients [*β*]), classification accuracy metrics (e.g. AUROC, C-index, AUPRC), but other relevant metrics (e.g. calibration [χ^2^]) can also be recorded. The covariates used in the model (most commonly age, sex, and genetic principal components (PCs) to account of population structure) are also linking to each set of metrics. Multiple PGS can be evaluated on the same Sample Set and further indexed as directly comparable **Performance Metrics**.

**Publications** provide provenance information for Scores and Performance Metrics (including those from external evaluations of existing PGS). Both journal articles and preprints can be indexed by either DOI or PubMed ID.

### The PGS Catalog: data content, access, and expansion

Any published or preprinted PGS can be added to the PGS Catalog provided it has (1) established analytic validity in external samples, and (2) the information necessary to calculate the score (see Supplemental Note 2 for additional details). To populate the PGS Catalog we screened over 180 publications for eligibility, of which 110 publications were eligible for curation and inclusion. The PGS Catalog currently contains 192 consistently-annotated PGS, curated from 69 publications (with the earliest published in 2008). These PGS predict a wide variety of diseases (e.g. cardiovascular diseases and different types of cancer) as well as anatomical (e.g. body mass index (BMI), bone density), cellular (e.g. blood cell counts and phenotypes) and molecular (serum urate, cholesterol and triglyceride levels) traits and measurements, encompassing 86 unique mapped ontology terms. To assess external validity the Catalog also indexes the results of evaluations of existing PGS in new contexts (e.g. direct comparisons of multiple PGS on the same sample); nine of these benchmarking publications evaluating nine existing PGS are also included in the current release of the PGS Catalog. Of the 68 publications developing at least one new PGS, nine also include a benchmarking of the performance to existing PGS.

The PGS Catalog can be accessed through a user interface (www.PGSCatalog.org) where indexed publications, scores and traits are browsable and searchable. Metadata describing PGS development and evaluation can be viewed on each score’s page (annotated example in Figure 1B). Pages describing traits with available PGS and the scores developed and evaluated within each publication can also be viewed (Supplemental Figure 1). Each PGS Scoring File contains a header describing the provenance of the score and consistently formatted columns describing the variants, alleles and weights. The Scoring File can be used in conjunction with common tools (e.g. PLINK^39^; (Supplemental Note 1)). The metadata and scoring files can be downloaded alone or in bulk from our website and FTP server; programmatic access to the database is also available through a RESTful API (complete implementation details are provided in Supplemental Note 3). Importantly the PGS Catalog provides users a source of existing scores that can be directly applied to their own data, making results obtained in PGS using the same score more comparable and circumventing the need to develop a new PGS for every application.

The Catalog identifies new papers from a manual literature search and user submissions, which subsequently undergo curation prior to their inclusion. Data curation and submission have been designed around a flexible template^40^, that allows common PGS development and evaluation details and results to be described according to our reporting items, and can be submitted directly to the Catalog for inclusion after validation by curators ^41^. Authors of PGS studies are encouraged to submit new PGS as well as subsequent PGS validations for indexing (by e-mail to), to grow the Catalog for the community, to maximize the utility of their PGS, and to enable reproducibility.

### Systematic evaluation of PGS yields comparable performance metrics

To demonstrate re-use and systematic comparison, we utilised the Catalog to assess the performance of nine PGSs for colorectal cancer in European, South Asian and African ancestries in the UK Biobank (UKB), a dataset external to all scores^42^ (methods described in Supplemental Note 4, cohort described in Supplemental Table 3). For each ancestry group, each PGS was evaluated using the standardised effect size of the PGS (OR/HR per standard deviation increase of PGS) and changes in classification accuracy (AUROC and C-index) as performance metrics (Figure 2, Supplemental Figure 2). Eight of the nine scores were predictive of colorectal cancer in European ancestries of UKB to varying degrees, and the magnitudes of effect sizes for two of the PGS were similar to that previously reported (Supplemental Figure 2). The score not significantly predictive of colorectal cancer in Europeans (PGS000151) comprised only 14 variants, and its predictive capacity in Europeans had not been previously evaluated. In South Asian and African ancestries of UKB, which combined are ~8% of total UKB individuals, the PGSs were largely not significantly predictive (Supplemental Table 2).

**Figure 2.**
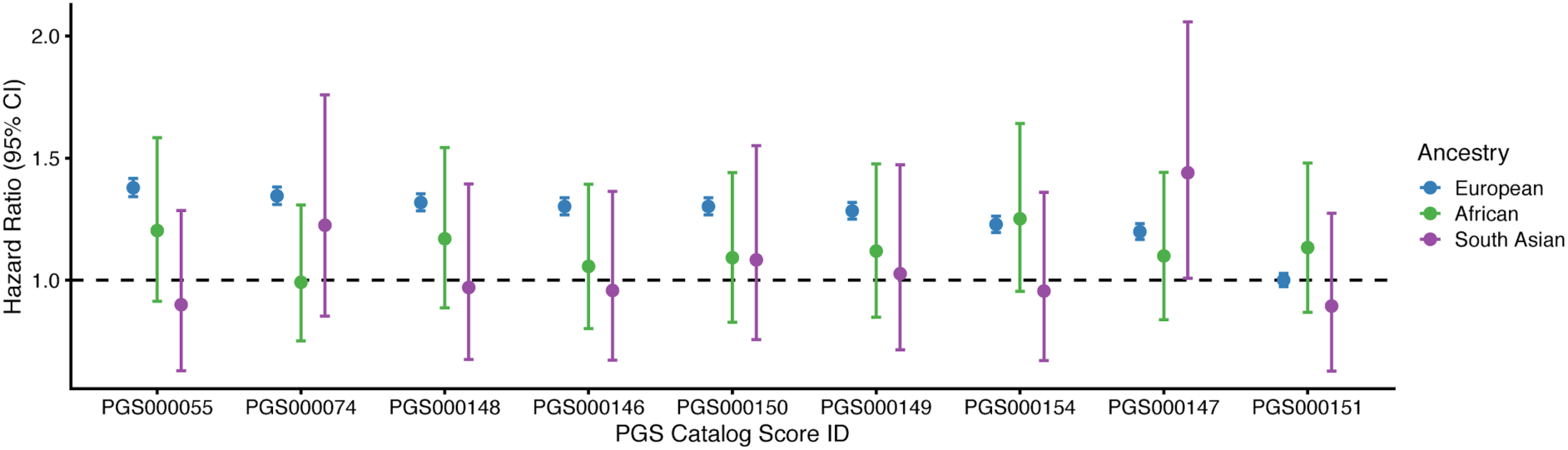
Benchmarking the association of nine colorectal cancer PGS in UKB. Each PRS was evaluated using a Cox proportional hazards regression model (age-as-timescale) to predict colorectal cancer status. Each model was fitting separately for each ancestry group. Standardised effect size (Hazard Ratio; HR), together with 95% confidence interval (CI), describes the increase in hazard per standard deviation increase of each PGS. Models were adjusted for sex, recruitment country, genotyping array, and the first 10 genetic principal components within each ancestry group.

### Conclusions and future developments

The PGS Catalog serves the community as a platform for polygenic score studies. The Catalog makes polygenic scores available for analysis in a standardised format along with consistent metadata, thereby enabling direct comparison between scores. We hope to facilitate reproducible PGS analyses by working with others towards standard formats and content of scoring files, and to provide new tools to support this (e.g. for validation and scoring). For instance, to address a common user request, we will harmonise PGS scoring files to frequently utilised genome builds (GRCh37 and 38). As the database grows, we will leverage the trait ontology to extend search functionality, allowing users to better identify and extract PGSs for any trait of interest.

PGS reproducibility must ensure that calculations are valid and consistent, with minimal variability across users. Based on community need, we intend to provide reference sample calculations and population distributions, similar to those for clinical tests. These enhancements will facilitate systematic and external PGS benchmarking studies, which are key to evaluating the validity of existing PGS.

As PGS increase in number, along with the diversity of phenotypes they predict, we will continue to grow the Catalog, curating new data and simplifying processes for researchers to deposit PGS they have developed and evaluated. We hope that researchers will join us in promoting data-sharing and submitting data so that the PGS Catalog provides a comprehensive resource for the community, enabling reproducibility as well as subsequent applications and translation of PGS.

## Data Availability

All data is available through the PGS Catalog (www.pgscatalog.org).

## Acknowledgments

We wish to thank all the authors of publications in the PGS Catalog for making their data available and indexable in our database, and wish to thank all those who responded to our inquiries and requests for data. This work makes use of UK Biobank Project #7439.This work was supported by core funding from: the UK Medical Research Council (MR/L003120/1), the British Heart Foundation (RG/13/13/30194;RG/18/13/33946) and the National Institute for Health Research [Cambridge Biomedical Research Centre at the Cambridge University Hospitals NHS Foundation Trust] [*]. This work was also supported by Health Data Research UK, which is funded by the UK Medical Research Council, Engineering and Physical Sciences Research Council, Economic and Social Research Council, Department of Health and Social Care (England), Chief Scientist Office of the Scottish Government Health and Social Care Directorates, Health and Social Care Research and Development Division (Welsh Government), Public Health Agency (Northern Ireland), British Heart Foundation and Wellcome. MI was supported by the Munz Chair of Cardiovascular Prediction and Prevention. This study was supported by the Victorian Government’s Operational Infrastructure Support (OIS) program. Research reported in this publication was supported by the National Human Genome Research Institute of the National Institutes of Health under Award Number U41HG007823. The content is solely the responsibility of the authors and does not necessarily represent the official views of the National Institutes of Health. In addition, we acknowledge funding from the European Molecular Biology Laboratory. JD holds a British Heart Foundation Chair and is funded by the National Institute for Health Research [Senior Investigator Award] [*]. MI and SR are supported by the National Institute for Health Research [Cambridge Biomedical Research Centre at the Cambridge University Hospitals NHS Foundation Trust].

*\*The views expressed are those of the authors and not necessarily those of the NHS, the NIHR or the Department of Health and Social Care*.

## Supplemental Note 1. PGS Catalog Scoring Files

The PGS Catalog’s Scoring File format is described on our website: https://www.pascataloa.org/downloads/. Each scoring file (variant information, effect alleles/weights) is formatted to be a gzipped tab-delimited text file, labelled by its PGS Catalog Score ID (e.g. PGS000001.txt.gz). We developed the scoring file format to closely resemble existing formats used to calculate scores in common software (e.g. PLINK) so that users could easily apply these scores within existing pipelines.

Scores are extracted from the relevant publication, and a consistent header (lines starting with #) has been added to each file listing relevant information about the PGS with links to the original publication and Catalog identifier:

### PGS CATALOG SCORING FILE - see www.pgscatalog.org/downloads/#dl_ftp for additional information ## POLYGENIC SCORE (PGS) INFORMATION # PGS ID = *PGS identifier, e.g. ‘PGS000001’* # Reported Trait = *trait, e.g. ‘Breast Cancer’* # Original Genome Build = *Genome build/assembly, e.g. ‘GRCh38’* # Number of Variants = *Number of variants listed in the PGS* ## SOURCE INFORMATION # PGP ID = *PGS publication identifier, e.g. ‘PGP000001’* # Citation = *Information about the publication*

rsID chr_name chr_position effect_allele reference_allele…

PGS scoring files are re-formatted to have consistent column headings based on the following schema:

| Column Header | Field Name | Field Description | Mandatory? |
| --- | --- | --- | --- |
| *rsID* | dbSNP Accession ID (rsID) | The SNP's rs ID | **YES** - Each PGS Scoring file must have either an *rsID* column or both a *chr_name* and *chr_position* column to identify the variant. |
| *chr_name* | Location - Chromosome | Chromosome name/number associated with the variant |  |
| *chr_position* | Location - Base pair position within the Chromosome | Chromosomal position associated with the variant |  |
| *effect_allele* | Effect Allele | The allele that's dosage is counted (e.g. {0, 1, 2}) and multiplied by the variant's weight ('effect_weight') when calculating score. The effect allele is also known as the 'risk allele'. | **YES** |
| *reference_allele* | Reference Allele | The other allele(s) at the loci | **Suggested** - most software requires this for the calculation of scores and matching of the variants to existing genotype data, |
| *effect_weight* | Variant Weight | Value of the effect that is multiplied by the dosage of the effect allele ('effect_allele') when calculating the score. | **YES** |
| *locus_name* | Locus Name | This is kept in for loci where the variant may be referenced by the gene (APOE e4). It is also common (usually in smaller PGS) to see the variants named according to the genes they impact. | *Optional* |
| *weight_type* | Type of Weight | Whether the author supplied Variant Weight is a: beta (effect size), or a log(OR/HR (odds/hazard ratio)) | *Optional* |
| *allelefrequency_effect* | Effect Allele Frequency | Reported effect allele frequency, if the associated locus is a haplotype then haplotype frequency will be extracted. | *Optional* |
| *is_interaction* | FLAG: Interaction | This is a TRUE/FALSE variable that flags whether the weight should be multiplied with the dosage of more than one variant. Interactions are demarcated with a *_x_* between entries for each of the variants present in the interaction. | *Optional* |
| *is_recessive* | FLAG: Recessive Inheritance Model | This is a TRUE/FALSE variable that flags whether the weight should be added to the PGS sum only if there are 2 copies of the effect allele (e.g. it is a recessive allele). | *Optional* |
| *is_haplotype* | FLAG: Haplotype or Diplotype | This is a TRUE/FALSE variable that flags whether the effect allele is a haplotype/diplotype rather than a single SNP. Constituent SNPs in the haplotype are semi-colon separated. | *Optional* |
| *is_diplotype* |  |  |  |
| *imputation_method* | Imputation Method | This describes whether the variant was specifically called with a specific imputation or variant calling method. This is mostly kept to describe HLA-genotyping methods (e.g. flag SNP2HLA, HLA*IMP) that gives alleles that are not referenced by genomic position. | *Optional* |
| *variant_description* | Variant Description | This field describes any extra information about the variant (e.g. how it is genotyped or scored) that cannot be captured by the other fields. | *Optional* |
| *inclusion_criteria* | Score Inclusion Criteria | Explanation of when this variant is included into the PGS (e.g. if it depends on the results from other variants). | Optional |

## Supplemental Note 2. Inclusion Criteria for the PGS Catalog

For the current PGS Catalog inclusion criteria see: https://www.pgscatalog.org/about/#eligibility. For a publication’s data to be included in the PGS Catalog, it must fulfill the following criteria for either a newly developed polygenic score or an evaluation of an existing score(s):

### A newly developed PGS

This includes the following information about the score and its predictive ability (evaluated on samples not used in training):

- Variant information necessary to apply the PGS to new samples (variant rsID and/or genomic position, weights/effect sizes, effect allele, genome build).
- Information about how the PGS was developed (computational method, variant selection, relevant parameters).
- Descriptions of the samples used for training (e.g. discovery of the variant associations [these can usually be extracted directly from the GWAS Catalog using GCST IDs], as well as fitting the PGS) and external evaluation.
- Establishment of the PGS’ analytic validity, and a description of its predictive performance (e.g. effect sizes [beta, OR, HR, etc.], classification accuracy, proportion of the variance explained (R2), and/or covariates evaluated in the PGS prediction).

### An evaluation of a previously developed PGS

This would include the evaluation of PGS already present in the Catalog (or one that meets the inclusion criteria specified above), on samples not used for PGS training. The requirements for description would be the same as for the evaluation of a new PGS.

## Supplemental Note 3. PGS Catalog Data Access and Implementation

Data in the PGS Catalog is provided under EMBL-EBI’s standard terms of use (https://www.ebi.ac.uk/about/terms-of-use/). The data in the Catalog can be currently accessed in the following three ways:

- **Bulk download** of the entire PGS Catalog’s metadata, describing all PGS in terms of their publication source, samples used for development/evaluation, and related performance metrics (details and links: www.pgscatalog.org/downloads/).
- The **PGS Catalog FTP server** (available at: https://ftp.ebi.ac.uk/pub/databases/spot/pgs/) is indexed by Polygenic Score (PGS) ID to allow programmatic access to the Scoring Files and metadata for each PGS, archived versions of the scoring files and metadata are also stored for reference (additional details: www.pgscatalog.org/downloads/).
- A **REST API** is also provided to allow programmatic access and querying of the PGS Catalog, better enabling other applications to be built on top of the resource. Endpoints to retrieve all or individual PGS Catalog data objects (Publications, Scores, Samples, Traits, Performance Metrics) are available (details at: https://www.pgscatalog.org/rest/).

The PGS Catalog is also is indexed on FAIRsharing.org (ref: bsg-d001448), and polygenic score identifiers (e.g. PGS000018) can be externally resolved via IDENTIFIERS.org (ref: pgs). A description of the FAIR indicators for the PGS Catalog are provided in Supplemental Table 1.

Additional bibliographic information for PGS Catalog **Publication** objects are retrieved from EuropePMC (e.g. title, authors, journal, publication dates)^43^. Additional information for each ontology term (e.g. synonyms, and mapped terms from other ontologies and disease coding resources [e.g. ICD/READ/SNOMED]) from the EFO ^35^ are obtained using the EMBL-EBI Ontology Lookup Service (OLS)^36^.

The PGS Catalog website and database are developed using the Django framework (version 3.0; https://djangoproject.com) in Python (version 3.7; https://www.python.org) with a PostgreSQL database (version 11; https://www.postgresql.org/). The website and database are both deployed on the Google Cloud (https://cloud.google.com/). The codebase for the Catalog can be viewed within our public GitHub repository (https://github.com/PGScatalog), currently provided under an Apache 2.0 License.

## Supplemental Note 4. Colorectal cancer benchmarking methods

To evaluate the predictive ability of PGS for colorectal cancer in the Catalog we used data from the UK Biobank (UKB), a cohort of ~500,000 participants from three countries (England, Wales, Scotland) of the United Kingdom^42^. Our analysis included 421,332 participants with genetic and phenotypic data (Supplemental Table 2), corresponding to 409,253 participants of European ancestry (UKB “White British” subset), 6,086 South Asian ancestry, and 5,984 African ancestry participants. South Asian (self-identifying as: Indian, Pakistani, or Bangladeshi) and African ancestry (self-identifying as: Caribbean, African, or Any other black background) participants were defined using an identical process to the White British participants, using principal components of genetic ancestry to identify a homogenous subset of self-identifying individuals by clustering^42^.

Diagnosis of colorectal cancer was performed using data linkage to the UK’s national cancer and death registries. Cases of colorectal cancer were identified using previously used ICD codes in UKB ^44^:

ICD9: 153.0 - 153.9, 154.0, 154.1, 154.8

ICD10: C18.0 - C18.9, C19, C20, C21.8

For each colorectal cancer diagnosis or death we recorded the date and age of the event. colorectal cancer events were defined as the first event of colorectal cancer, and participants were censored after the last cancer registry linkage date (2016-03-31). We excluded 449 participants who had self-reported history of colorectal cancer at recruitment and no linked cancer registry data.

PGS files were downloaded from the PGS Catalog and scores for each participant were calculated using PLINK^39^. Scores were standardised within each ancestry; the mean and standard deviation for colorectal cancer cases and controls are reported by ancestry group (Supplemental Table 3).

Each score’s predictive ability is measured in terms of classification of cases vs controls, via the standardised effect size of the PGS (OR/HR per standard deviation increase of PGS) and classification accuracy (AUROC and concordance statistic [C-index]). We measured the HR and C-index using a Cox Proportional Hazards model with age-as-timescale, adjusting for sex, genotyping array, country of recruitment, and 10 PCs of genetic ancestry. We measured the OR and AUROC using a logistic regression model adjusting for the sex, age at recruitment, country of recruitment, genotyping array, and 10 PCs of genetic ancestry. The effect sizes are reported with the 95% confidence interval for each PGS (Supplemental Table 3). Statistical analyses were performed in python: the Cox model was implemented using the *lifelines* package^45^, and logistic regression was performed using the *statsmodels* package^46^.

## Supplemental Figure

**Supplemental Figure 1.**
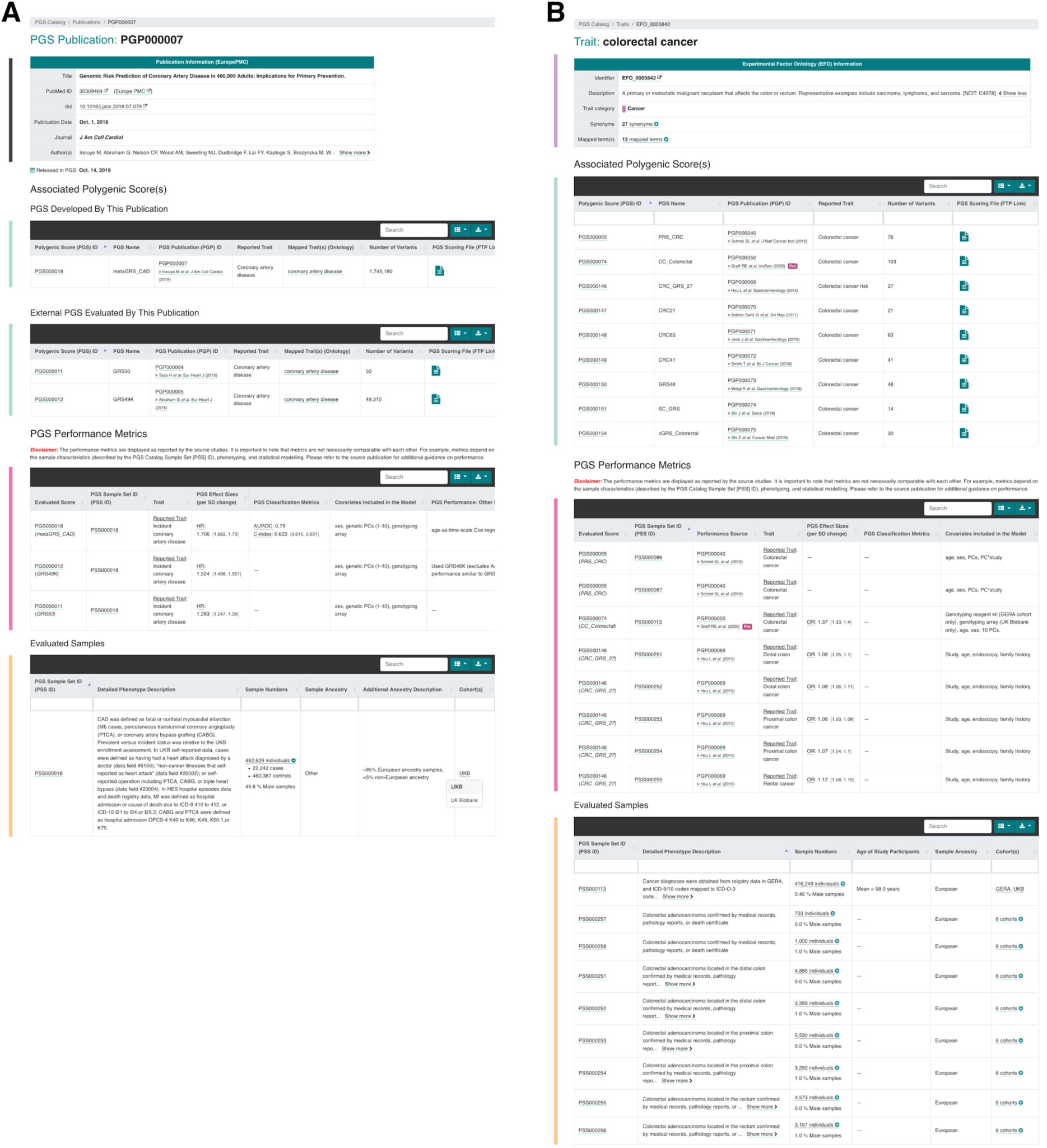
Examples of PGS Catalog Publication and Trait website pages. (**A**) Example of how each Publication and its related metadata (links to publication, EuropePMC, and PGS that were developed and evaluated within the paper) are displayed on PGSCatalog.org (example publication PGP00007^12^). (**B**) Example of how each Trait (ontology term, description, synonyms, and mapped terms [e.g. ICD/SNOMED] extracted from EFO^35,36^) and its related metadata (PGS that have predicted the current trait, and subsequent evaluation of those scores) are displayed on PGSCatalog.org (example trait: colorectal cancer, EFO_0005842). Sections of each webpage are highlighted with coloured bars corresponding to the data objects they display in **Figure 1A**.

**Supplemental Figure 2.**
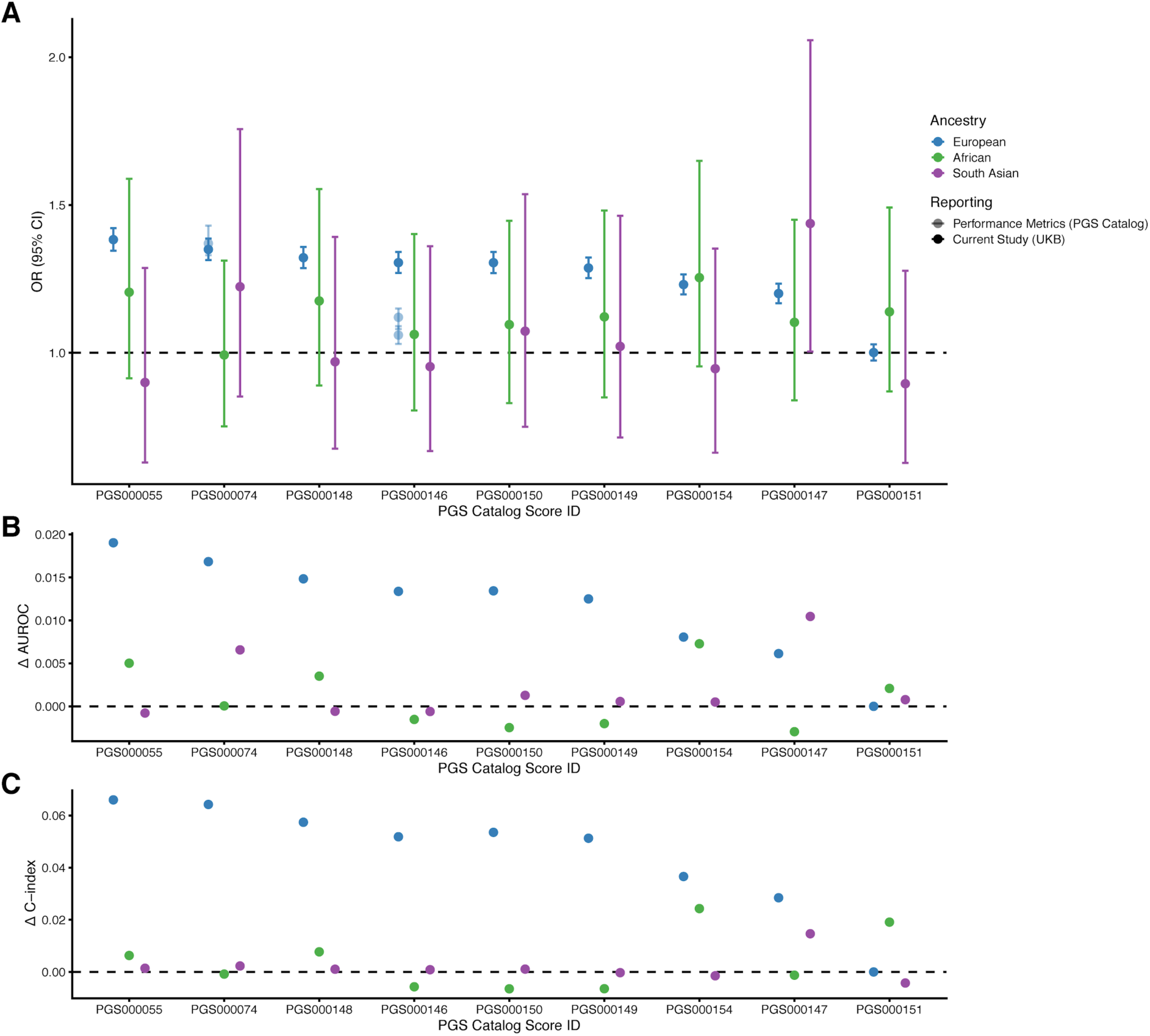
Performance Metrics for colorectal cancer PGS in UKB. Each PRS was evaluated within a logistic regression model for predicting colorectal cancer status for participants in UKB (**A-B**), and a separate Cox proportional hazards regression model (age-as-timescale) (**Figure 2, C**). (**A**) Standardised effect size (Odds Ratio; OR) describing the odds of having colorectal cancer per unit increase in each PGS. Previously reported effect sizes that were recorded in the Catalog are also plotted for PGS000074 and PGS000146. (**B**) Change in model classification accuracy (Area Under the Reciever Operating Characteristic Curve; AUROC) when the PGS is added to a logistic regression model including the existing covariates (age at recruitment, sex, recruitment country, genotyping array, and 10 PCs of genetic ancestry). (**C**) Change in model classification accuracy (concordance statistic; C-index) when the PGS is added to a risk model including the existing covariates (sex, recruitment country, genotyping array, and 10 principal components [PCs] of genetic ancestry).

## Supplemental Tables

**Supplemental Table 1.** FAIR indicators of PGS Catalog. This table describes details of how the current PGS Catalog conforms to FAIR data principles. For the purposes of this table the Score constitutes the data (e.g. variants, effect weights and alleles), and is linked to metadata (Samples, Performance Metrics, Publications) describing it.

| Core FAIR principle | FAIR principle | PGS Catalog indicator |
| --- | --- | --- |
| **Findable** | F1. (meta)data are assigned a globally unique and persistent identifier | Each polygenic score is assigned a unique identifier (e.g. PGS000018) that is linked to all relevant metadata and publication sources in the Catalog. The PGS identifier can be resolved externally through [IDENTIFIERS.org](https://identifiers.org) (prefix: *pgs*) |
|  | F2. data are described with rich metadata (defined by R1 below) | Polygenic scores included in the database are well-described, both in terms of their provenance and ability to be applied. Details in Supplemental Table 1 and on our website at: http://www.pgscatalog.org/docs/ |
|  | F3. metadata clearly and explicitly include the identifier of the data it describes | All metadata is linked to either a Polygenic Score (PGS), Sample Set (PSS), Performance Metric (PPM), or Publication (PGP) ID within the database. Ontology terms are described using the identifiers from the Experimental Factor Ontology. Publication sources are described using DOI and PMID.<br>Scoring files for each PGS are labelled with their PGS ID, and findable with the metadata on our FTP (http://ftp.ebi.ac.uk/pub/databases/spot/pgs/) described here: http://www.pgscatalog.org/downloads/ |
|  | F4. (meta)data are registered or indexed in a searchable resource | The PGS Catalog is indexed at [FAIRsharing.org](https://fairsharing.org) (ID: *bsg-d001448*) and indexed by Google Search. |
| **Accessible** | A1. (meta)data are retrievable by their identifier using a standardized communications protocol | Metadata can be easily viewed on our web interface (www.pgscatalog.org) with visible download links for each Score.<br>Scoring files and metadata can also be browsed and downloaded from our FTP site by PGS ID.<br>The full Catalog can also be accessed using our REST API: https://www.pgscatalog.org/rest/. |
|  | A1.1 the protocol is open, free, and universally implementable | Yes, the www.pgscatalog.org website is freely accessible to all. |
|  | A1.2 the protocol allows for an authentication and authorization procedure, where necessary | Not applicable |
|  | A2. metadata are accessible, even when the data are no longer available | Archived versions of the scoring files and metadata are stored for the complete database as well as individual scores on our FTP (http://ftp.ebi.ac.uk/pub/databases/spot/pgs/) |
| **Interoperable** | I1. (meta)data use a formal, accessible, shared, and broadly applicable language for knowledge representation. | PGS metadata is distributed from our API using JSON formats, the REST API is documented using the OpenAPI Specification (OAS3; https://github.com/OAI/OpenAPI-Specification/blob/master/versions/3.0.2.md). |
|  | I2: (Meta)data use vocabularies that follow the FAIR principles | The PGS identifier can be resolved externally through IDENTIFIERS.org (prefix: *pgs*) |
|  | I3. (meta)data include qualified references to other (meta)data | Traits are represented using (represented using ontology terms) associated with PGS are linked to the Experimental Factor Ontology (EFO) terms and include links to the EFO. |
| **Reusable** | R1. meta(data) are richly described with a plurality of accurate and relevant attributes | Polygenic scores included in the database are well-described, both in terms of their provenance and ability to be applied. Details in Supplemental Table 1 and on our website at: http://www.pgscatalog.org/docs/ |
|  | R1.1. (meta)data are released with a clear and accessible data usage license | All data are made available through EMBL-EBI's standard terms of use (https://www.ebi.ac.uk/about/terms-of-use/) |
|  | R1.2. (meta)data are associated with detailed provenance | Each PGS and Performance Metric is linked to a source Publication that can be accessed by either a digital object identifier (DOI) or PubMed ID (PMID). |
|  | R1.3. (meta)data meet domain-relevant community standards | The PGS Catalog is consistent with Polygenic Risk Score Reporting Standards (PRS-RS) <sup>24</sup> |

**Supplemental Table 2.**
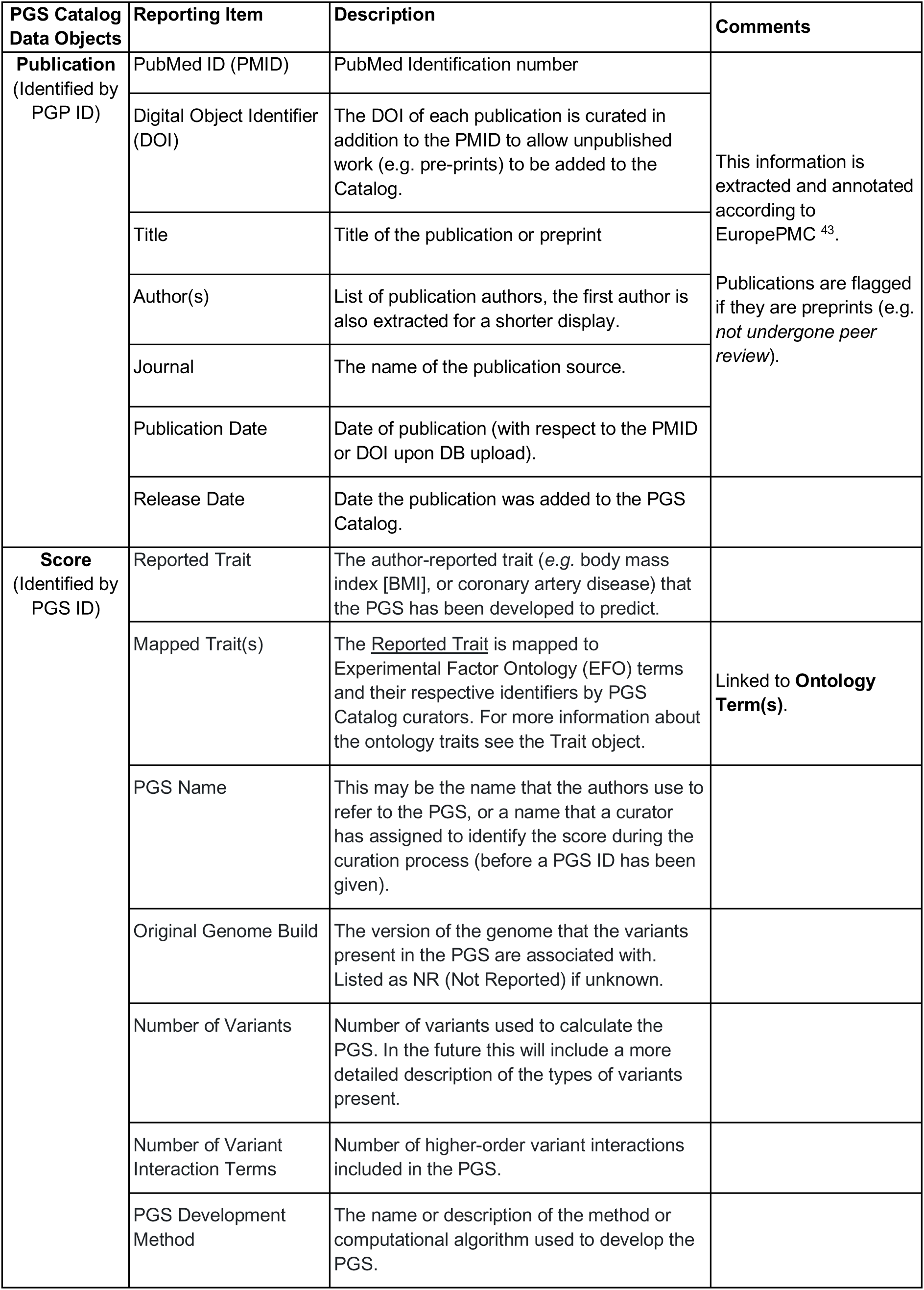
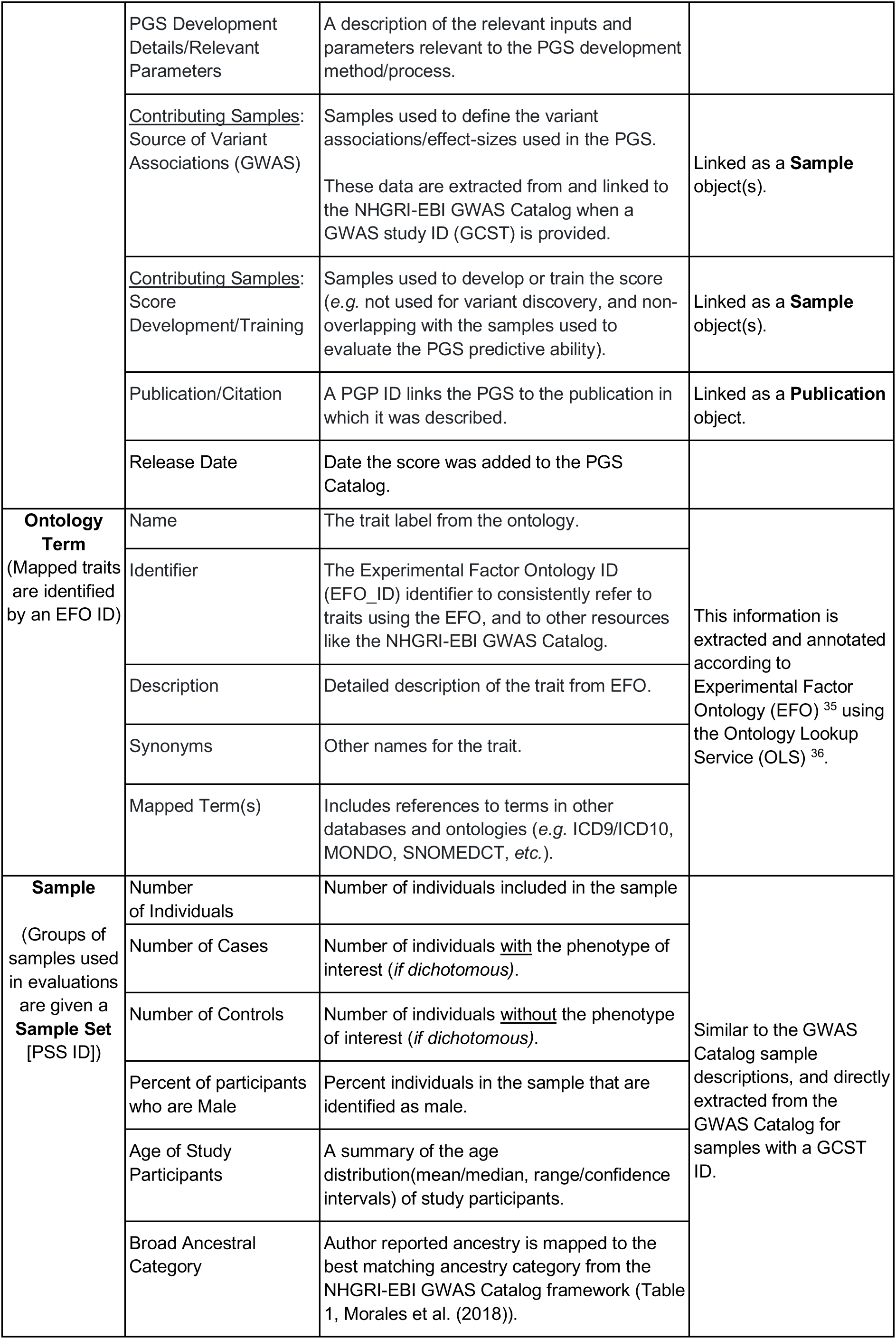
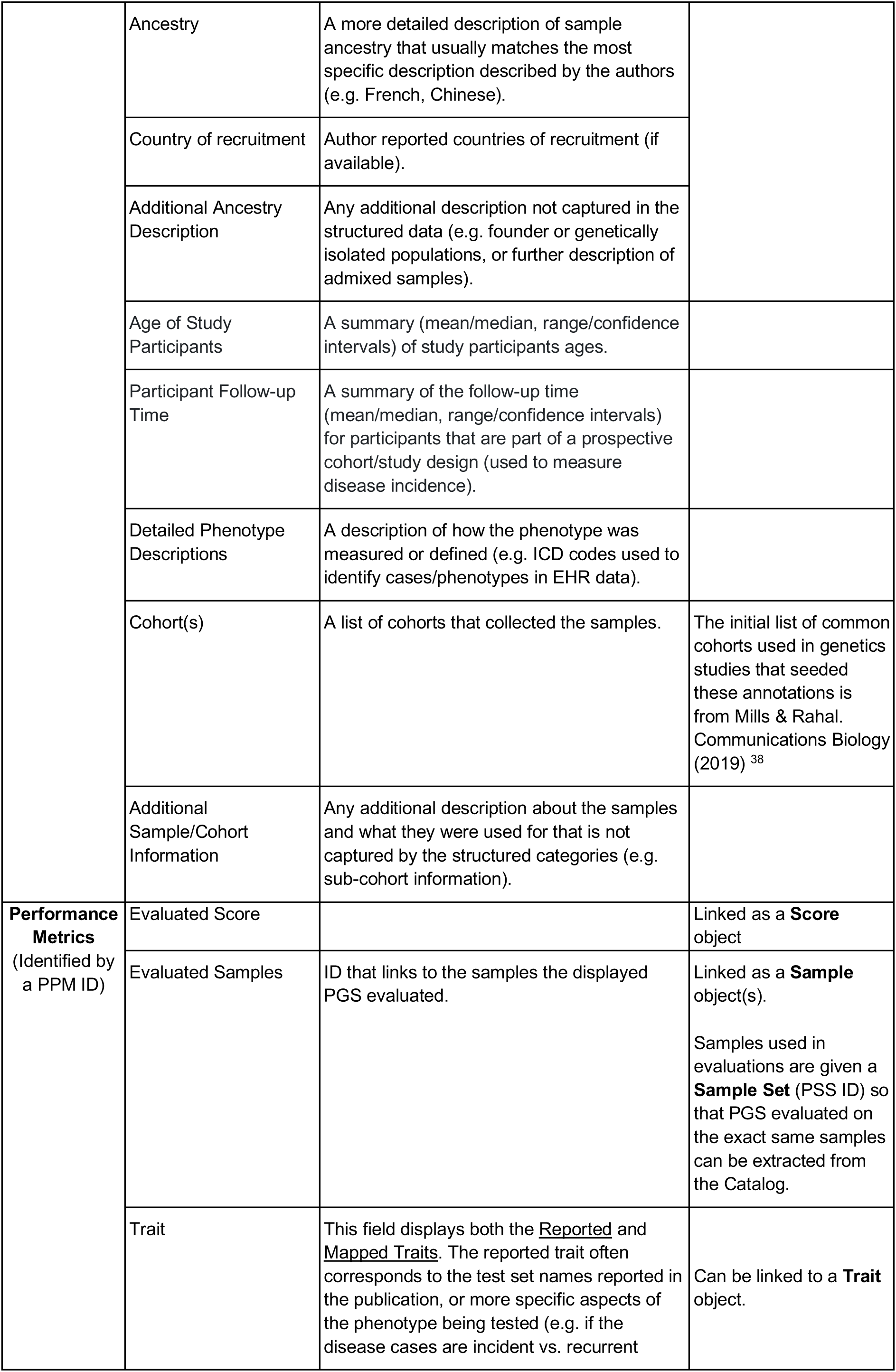
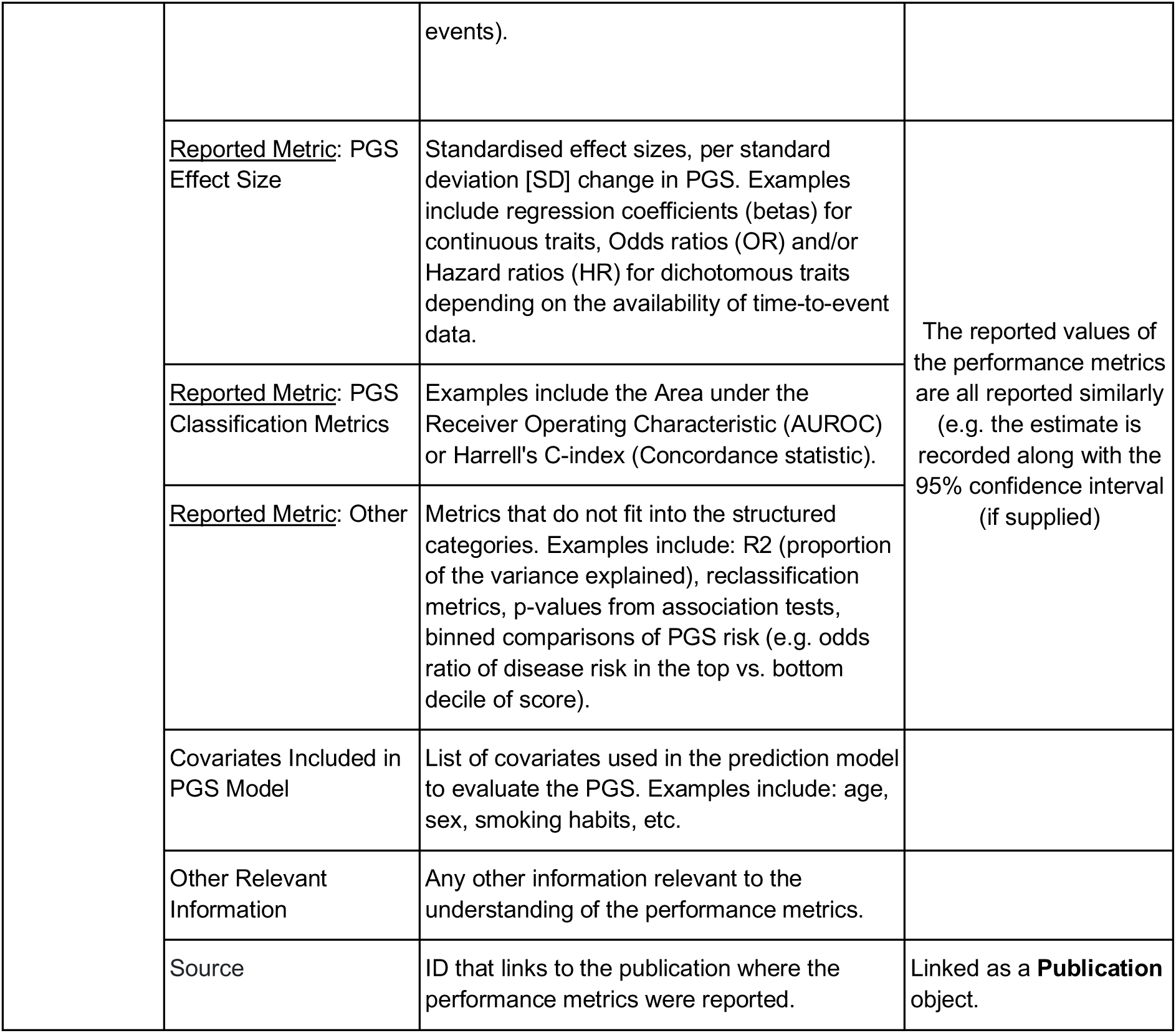
PGS Catalog Reporting Items. This table describes the reporting items that can be captured for each of the data objects in the PGS Catalog.

| PGS Catalog Data Objects | Reporting Item | Description | Comments |
| --- | --- | --- | --- |
| **Publication**<br>(Identified by PGP ID) | PubMed ID (PMID) | PubMed Identification number | This information is extracted and annotated according to EuropePMC <sup>43</sup>.<br>Publications are flagged if they are preprints (e.g. *not undergone peer review*). |
|  | Digital Object Identifier (DOI) | The DOI of each publication is curated in addition to the PMID to allow unpublished work (e.g. pre-prints) to be added to the Catalog. |  |
|  | Title | Title of the publication or preprint |  |
|  | Author(s) | List of publication authors, the first author is also extracted for a shorter display. |  |
|  | Journal | The name of the publication source. |  |
|  | Publication Date | Date of publication (with respect to the PMID or DOI upon DB upload). |  |
|  | Release Date | Date the publication was added to the PGS Catalog. |  |
| **Score**<br>(Identified by PGS ID) | Reported Trait | The author-reported trait (e.g. body mass index [BMI], or coronary artery disease) that the PGS has been developed to predict. |  |
|  | Mapped Trait(s) | The <u>Reported Trait</u> is mapped to Experimental Factor Ontology (EFO) terms and their respective identifiers by PGS Catalog curators. For more information about the ontology traits see the Trait object. | Linked to **Ontology Term(s)**. |
|  | PGS Name | This may be the name that the authors use to refer to the PGS, or a name that a curator has assigned to identify the score during the curation process (before a PGS ID has been given). |  |
|  | Original Genome Build | The version of the genome that the variants present in the PGS are associated with. Listed as NR (Not Reported) if unknown. |  |
|  | Number of Variants | Number of variants used to calculate the PGS. In the future this will include a more detailed description of the types of variants present. |  |
|  | Number of Variant Interaction Terms | Number of higher-order variant interactions included in the PGS. |  |
|  | PGS Development Method | The name or description of the method or computational algorithm used to develop the PGS. |  |
|  | PGS Development Details/Relevant Parameters | A description of the relevant inputs and parameters relevant to the PGS development method/process. |  |
|  | <u>Contributing Samples:</u><br>Source of Variant Associations (GWAS) | Samples used to define the variant associations/effect-sizes used in the PGS.<br>These data are extracted from and linked to the NHGRI-EBI GWAS Catalog when a GWAS study ID (GCST) is provided. | Linked as a **Sample** object(s). |
|  | <u>Contributing Samples:</u><br>Score Development/Training | Samples used to develop or train the score (e.g. not used for variant discovery, and non-overlapping with the samples used to evaluate the PGS predictive ability). | Linked as a **Sample** object(s). |
|  | Publication/Citation | A PGP ID links the PGS to the publication in which it was described. | Linked as a **Publication** object. |
|  | Release Date | Date the score was added to the PGS Catalog. |  |
| **Ontology Term**<br>(Mapped traits are identified by an EFO ID) | Name | The trait label from the ontology. | This information is extracted and annotated according to Experimental Factor Ontology (EFO) <sup>35</sup> using the Ontology Lookup Service (OLS) <sup>36</sup>. |
|  | Identifier | The Experimental Factor Ontology ID (EFO_ID) identifier to consistently refer to traits using the EFO, and to other resources like the NHGRI-EBI GWAS Catalog. |  |
|  | Description | Detailed description of the trait from EFO. |  |
|  | Synonyms | Other names for the trait. |  |
|  | Mapped Term(s) | Includes references to terms in other databases and ontologies (e.g. ICD9/ICD10, MONDO, SNOMEDCT, etc.). |  |
| **Sample**<br>(Groups of samples used in evaluations are given a **Sample Set** [PSS ID]) | Number of Individuals | Number of individuals included in the sample | Similar to the GWAS Catalog sample descriptions, and directly extracted from the GWAS Catalog for samples with a GCST ID. |
|  | Number of Cases | Number of individuals <u>with</u> the phenotype of interest (*if dichotomous*). |  |
|  | Number of Controls | Number of individuals <u>without</u> the phenotype of interest (*if dichotomous*). |  |
|  | Percent of participants who are Male | Percent individuals in the sample that are identified as male. |  |
|  | Age of Study Participants | A summary of the age distribution(mean/median, range/confidence intervals) of study participants. |  |
|  | Broad Ancestral Category | Author reported ancestry is mapped to the best matching ancestry category from the NHGRI-EBI GWAS Catalog framework (Table 1, Morales et al. (2018)). |  |
|  | Ancestry | A more detailed description of sample ancestry that usually matches the most specific description described by the authors (e.g. French, Chinese). |  |
|  | Country of recruitment | Author reported countries of recruitment (if available). |  |
|  | Additional Ancestry Description | Any additional description not captured in the structured data (e.g. founder or genetically isolated populations, or further description of admixed samples). |  |
|  | Age of Study Participants | A summary (mean/median, range/confidence intervals) of study participants ages. |  |
|  | Participant Follow-up Time | A summary of the follow-up time (mean/median, range/confidence intervals) for participants that are part of a prospective cohort/study design (used to measure disease incidence). |  |
|  | Detailed Phenotype Descriptions | A description of how the phenotype was measured or defined (e.g. ICD codes used to identify cases/phenotypes in EHR data). |  |
|  | Cohort(s) | A list of cohorts that collected the samples. | The initial list of common cohorts used in genetics studies that seeded these annotations is from Mills & Rahal. Communications Biology (2019) <sup>38</sup> |
|  | Additional Sample/Cohort Information | Any additional description about the samples and what they were used for that is not captured by the structured categories (e.g. sub-cohort information). |  |
| **Performance Metrics**<br>(Identified by a PPM ID) | Evaluated Score |  | Linked as a **Score** object |
|  | Evaluated Samples | ID that links to the samples the displayed PGS evaluated. | Linked as a **Sample** object(s).<br>Samples used in evaluations are given a **Sample Set** (PSS ID) so that PGS evaluated on the exact same samples can be extracted from the Catalog. |
|  | Trait | This field displays both the <u>Reported</u> and <u>Mapped Traits</u>. The reported trait often corresponds to the test set names reported in the publication, or more specific aspects of the phenotype being tested (e.g. if the disease cases are incident vs. recurrent events). | Can be linked to a **Trait** object. |
|  | <u>Reported Metric:</u> PGS Effect Size | Standardised effect sizes, per standard deviation [SD] change in PGS. Examples include regression coefficients (betas) for continuous traits, Odds ratios (OR) and/or Hazard ratios (HR) for dichotomous traits depending on the availability of time-to-event data. | The reported values of the performance metrics are all reported similarly (e.g. the estimate is recorded along with the 95% confidence interval (if supplied)) |
|  | <u>Reported Metric:</u> PGS Classification Metrics | Examples include the Area under the Receiver Operating Characteristic (AUROC) or Harrell's C-index (Concordance statistic). |  |
|  | <u>Reported Metric:</u> Other | Metrics that do not fit into the structured categories. Examples include: R2 (proportion of the variance explained), reclassification metrics, p-values from association tests, binned comparisons of PGS risk (e.g. odds ratio of disease risk in the top vs. bottom decile of score). |  |
|  | Covariates Included in PGS Model | List of covariates used in the prediction model to evaluate the PGS. Examples include: age, sex, smoking habits, etc. |  |
|  | Other Relevant Information | Any other information relevant to the understanding of the performance metrics. |  |
|  | Source | ID that links to the publication where the performance metrics were reported. | Linked as a **Publication** object. |

**Supplemental Table 3.**
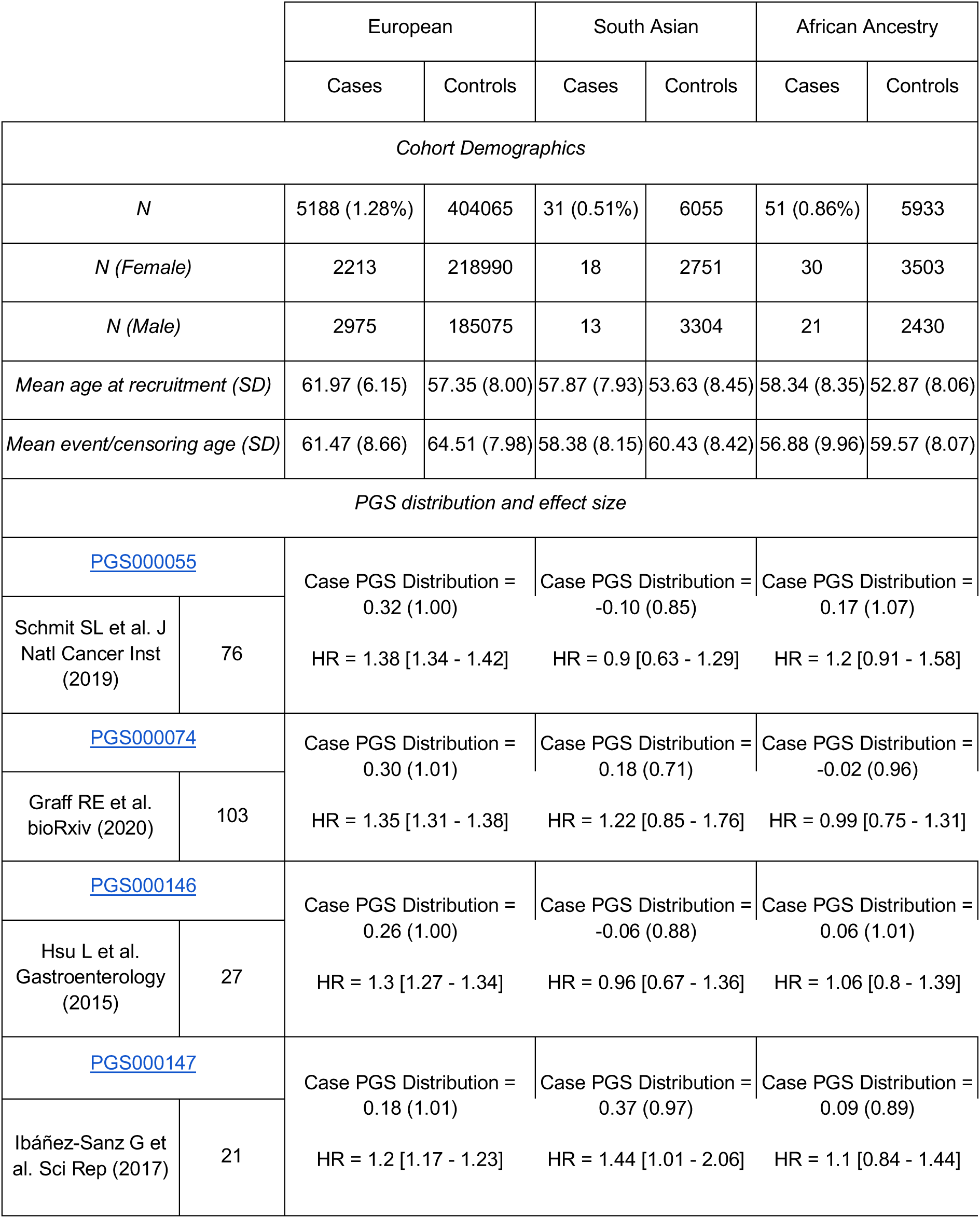
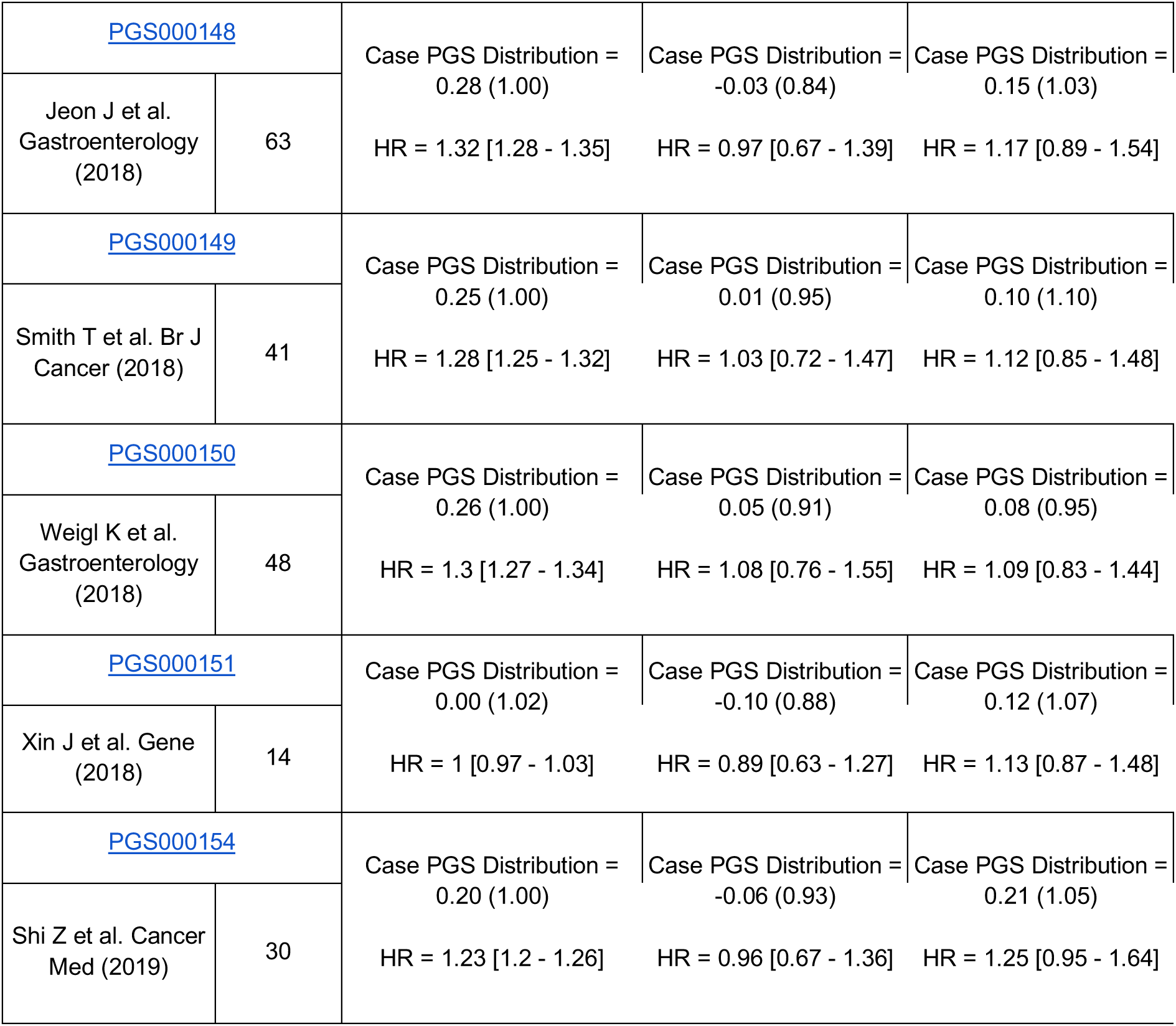
UKB Benchmarking cohort description and results. Cohort age and sex demographics broken down by colorectal cancer case/control status and participant ancestry. The distribution (mean and standard deviation [SD]) of each standardised PGS in colorectal cancer cases is also given, along with its effect size (Hazard Ratio; HR), citation and number of variants included in the PGS; the distribution of each PGS in controls is zero-mean and unit-variance.

